# Moving from Bench to Bedside: A Social History of Actionability in Biomedicine

**DOI:** 10.64898/2026.08.03.26359586

**Authors:** Kellie Owens, Ari Z. Klein, Graciela Gonzalez-Hernandez

## Abstract

**Background:** As medicine grows increasingly technological and scientific, biomedical researchers working to bring new knowledge to bear on clinical practice face a key question: when is a new intervention or treatment ready for clinical use? Because this is both a technical and ethical dilemma, it is crucial to examine the social history of how different approaches to this question emerge and the values or assumptions they embed.

**Methods:** We examine the rise and proliferation of an increasingly common framework for assessing the value of new biomedical data or technology, “actionability,” through a computational analysis of published scientific literature referencing this and related terms, including topic modeling and Medical Subject Headings (MeSH) term analysis of over 7000 scientific abstracts indexed in PubMed.

**Results:** We find that actionability, as a term, began appearing more commonly in published literature in the mid-2000s, and proliferated throughout the 2010s and into the 20s. While originally used primarily in research on healthcare quality and implementation, the concept’s rise in popularity is ultimately driven by uptake in the fields of clinical genetics and oncology.

**Conclusions:** The adoption of actionability in these fields suggests that actionability as a conceptual framework may be most valuable to areas of translational medicine seeking to make sense of increasing amounts of data and technological innovation with differing levels of scientific validity and clinical utility. Recognizing this value, we also caution that actionability drives our attention primarily towards whether a test or piece of information can lead to action, not whether that action has proven benefits. As clinicians and researchers face difficult questions about how to sort through growing amounts of data to generate knowledge that can have a real impact on patient health, empirical bioethics should play a key role in analyzing the trade-offs and impacts of different approaches.

## Introduction

Over the course of the twentieth century, biological sciences like genetics and biochemistry have reshaped clinical understandings of disease mechanisms, screening techniques, and treatments. Because medicine grew increasingly technological and scientific, its practice became more complex, multi-sited, and multidirectional—a process social scientists have called “biomedicalization” (Clarke et al. 2003). At the same time, generating medical evidence similarly requires more standardized approaches and collective action among a range of actors and organizations (Cambrosio et al. 2006). Over the course of this shift, biomedical researchers working to bring new scientific knowledge to bear on clinical medicine face a complicated question: when is a new technology or treatment ready to use with patients? This is both a technical and ethical dilemma that researchers and clinicians approach in many ways. For example, researchers developed concepts like “clinical validity:” what is the predictive value of a test for a clinical outcome, such as, how likely is cancer to occur in a patient with a positive test? Or, the concept of “clinical utility:” how likely is it for a test to cause an improved health outcome (National Cancer Institute)?

This article traces the social history of another related concept that has become a dominant paradigm for considering the translation of biomedical research into clinical care: “actionability.” Actionability directs attention toward whether new data or technological innovation warrants action and reflects an urgency towards communicating information that may provide clinicians or patients with immediate or future benefit. Actionability is often used to discuss what counts as a clinically-relevant test, which types of results are worthy of returning to patients, and what actions, if any, should be taken after a patient receives a test result. It is also used to discuss how information generated from a test can increase or decrease medical uncertainty. While concepts like actionability often have technical definitions, their interpretation and implementation differs based on social context.

Actionability is more than just a concept used in the literature; it is practiced and structures decision-making in a variety of ways. For example, Stivers and Timmermans (2017) find that genetic testing for children with disabilities becomes “actionable,” even if it does not change any clinical care, because it helps sort through or solve other problems and needs of families raising these children.

To date, there is little analysis of the meaning, emergence, and proliferation of actionability as a framework for translational research and practice. This is a critical gap because the values and assumptions embedded within discussions of actionability differ from other regimes of knowledge production and practice in translational medicine. To illuminate the emergence of this framework and its embedded meanings and assumptions, we utilize computational methods such as topic modeling to trace how researchers use and understand the value of this concept in published literature. This approach, in conjunction with our prior published qualitative research on the politics of actionability (Owens 2021), provides a more comprehensive examination of how actionability as a translational framework organizes biomedical science and clinical care.

We find that actionability, as a term, began appearing more commonly in published literature in the mid-2000s, and proliferated throughout the 2010s and into the 20s. While originally used primarily in research on health care quality and implementation, the concept’s rise in popularity is ultimately driven by uptake in the fields of clinical genetics and oncology. The uptake of actionability in these fields suggests that actionability as a conceptual framework may be most valuable to areas of translational medicine seeking to make sense of increasing amounts of data and technological innovation with differing levels of scientific validity and clinical utility.

### The politics of evidence and efficacy

Interrogating the framework of actionability requires situating this concept within larger transformations of medicine and science over the course of the twentieth century. Social scientists employ the term “biomedicine” to capture the complex intertwining of the biological and clinical in modern health care science and practice. The production of clinical knowledge is no longer limited to examination rooms, and the generation of biotechnological knowledge extends beyond laboratories. In practical terms, laboratory research and healthcare practices are now more closely interconnected than ever before. As medicine has become increasingly technoscientific, there has been a deliberate focus on translational research and “learning healthcare systems” to bridge the gap between research and practice by consistently integrating new biological data into clinical care.

This transformation necessitates the adoption of collective and coordinated models of objectivity, as evidenced by existing research on how the consolidation of standard procedures and regulations gets negotiated (Cambrosio et al. 2006). The generation of biomedical evidence now demands new practices such as inter-laboratory agreements, multi-center clinical trials, consensus workshops, and the establishment of biobanks and interoperable datasets between healthcare systems. Biomedicine has also taken on new organizational forms, forging connections between academic laboratories and profit-driven companies while simultaneously reshaping disciplinary boundaries.

These new arrangements simultaneously require new ways of thinking about what counts as clinical evidence and which types of clinical evidence warrant actions. With the rise of the evidence-based medicine (EBM) movement in the 1990s, biomedicine began creating standardized tools to assess the utility of medical evidence and interventions and paid greater attention to the harms of acting on ineffective technology (Timmermans and Berg 2010). The EBM movement generated a hierarchy of clinical evidence, placing the randomized clinical trial (RCT) or meta-analyses of many RCTs at the top of the pyramid. At the same time, these types of clinical trials are expensive and difficult to complete. In some fields, researchers and clinicians may feel a moral obligation to offer patients potentially beneficial information or treatments before clinical trials can be completed (Frieden 2017).

As social scientists have previously demonstrated, there are, in practice, many “scientific methods” that researchers deploy in the generation of new scientific evidence (Knorr Cetina 1999). In some types of biomedical research, scientists develop a hypothesis or set of research questions and design their data collection and analysis with those particular questions in mind. But as we enter the era of “big data,” a new and competing research paradigm has emerged: collect as much data as possible, without specific goals in mind, and generate findings based on subsequent analyses of that data. For example, the NIH All of Us research program is designed to collect a wide range of clinical and social data from one million participants, to be stored in a biobank for researchers to study over the course of decades (Denny and Collins 2021). As researchers comb through this data, they face a more immediate ethical challenge than may be present in other types of research with clearer beginning and end points: when are their study findings certain and important enough that they might have a duty to return results to current or future patients? The following section presents a brief analysis of different frameworks that have emerged to address this question.

### On “actionability,” “validity,” and “utility”

The rise of “actionability” in published biomedical literature rests on the foundations of a range of concepts that assess value in translational medicine. For example, around the completion of the Human Genome Project in 2003, the Centers for Disease Control (CDC) Office of Public Health Genomics developed the “ACCE Model Project” for evaluating genetic tests (Centers for Disease Control 2010). This approach included analytic validity, clinical validity, clinical validity, and associated ethics concerns as key elements in assessing the value of a genetic test. Analytic validity refers to the ability of a test to measure the property or characteristic it was designed to measure. Clinical validity refers to how consistently and accurately a tool can predict a given outcome. Clinical utility is a broader measure of whether a tool improves relevant clinical outcomes. As bioethics researchers have demonstrated, the concept of “utility” is not new, and is conceptualized and assessed in different ways depending on the field and context. Utility may refer to a balance of benefits and harms, to usefulness in personal or clinical scenarios, or be used as a metric of health-related quality of life (Smith et al. 2021).

Given that a number of concepts already existed to assess value in medicine, why did actionability arise as a new and competing framework? As other social scientists have documented, the rise of translational science leads to the problem of identifying what counts as an “actionable outcome” (Nelson et al. 2013). Because actionability embeds different values and assumptions than other frameworks, it is crucial to understand what we gain and lose when this framework emerges and gains traction. To begin answering this question, our prior qualitative research on actionability documented its meaning within clinical genetics, finding that clinicians often referenced actionability in the context of two main assessments of: (1) the quality and quantity of evidence for a tool or test, and (2) perceived capacity and infrastructural barriers to implementation (Owens 2021). Prior research has also shown that interpretations of actionability are highly variable, relying as much on social context as scientific theory. Professional societies, such as the American College of Medical Genetics and Genomics (ACMG) and the European Society for Human Genetics, disagree in their conceptualizations of actionability, and the ACMG has changed the ethical underpinnings of its related guidance on the reporting of “actionable” genetic findings multiple times (Owens 2021). Patients also have nuanced, variable understandings of actionability that do not always match expert definitions of actionability (Facio et al. 2012; Jamal et al. 2017; Gornick 2018). For example, patients are more likely than clinicians to view genomic information as actionable if it could foster lifestyle changes or differences in reproductive decision-making (Lewis et al. 2015; Mackley et al. 2017).

In practice, the concept of actionability is thus highly contextual and incorporates features of many other concepts, including analytic validity, clinical validity, and clinical utility. This flexibility may be the framework’s greatest strength, and may be why it is more valuable to some fields than others. To more deeply explore which fields have taken up this concept, and understand the rise and proliferation of actionability in biomedicine more generally, we use a variety of computational methods to analyze published literature on this topic.

## Methods

### Data collection and validation

First, we sought to collect systematic data about published scientific articles referencing actionability. In December 2022, we used the PubMed application programming interface (API) to search for abstracts that contained the word “actionable” or “actionability,” which returned 8943 abstracts. For each abstract, we retrieved the text, PubMed identifier (PMID), year of publication, and Medical Subject Headings (MeSH) terms. MeSH terms are standardized keywords applied to articles manually by the National Library of Medicine (NLM) to provide a standardized index of an article’s content (National Library of Medicine 2023). Of the 8943 abstracts, 7074 (79%) included MeSH terms. Despite conducting our search in 2022, 20 abstracts were indexed with 2023 publication dates; however, these abstracts do not constitute a full analysis of data from 2023. We used the PyMed Python library to clean the abstracts, removing metadata and markup for human readability and downstream automated processing. To assess the extent to which the abstracts were within the scope of this study, an author and a research assistant independently reviewed a random sample of 100 abstracts and labeled them for whether or not they were broadly related to biomedicine, determining that 95% of them were, with 100% agreement between the authors. We considered this proportion of relevant abstracts to be sufficient for aggregate analysis, deciding not to review all 8943 abstracts for inclusion/exclusion.

### Topic modeling

In addition to analyzing the MeSH terms associated with the abstracts, we used an unsupervised machine learning method called *topic modeling* to discover groups of words that frequently co-occur in the abstracts themselves. We generated these groups of words, or topics, using a statistical technique called *Latent Dirichlet Allocation (LDA)*, which treats each abstract as a composite of multiple topics. Prior to generating the topics, we preprocessed the abstracts by lemmatizing the words. Lemmatization converts a word to its root form so that different words with the same root form–for example, *patient* and *patients*– can be semantically analyzed as a single word in a topic. We used the WordNet Lemmatizer in the Natural Language Toolkit (NLTK) Python library. In addition, we removed stopwords–common words that do not contribute substantive meaning, such as articles, conjunctions, prepositions, and pronouns–and other words that would be uninformative for interpreting the topics in this study, including “actionable” and “actionability.”

To optimize the topic model, we experimented with tuning the number of training iterations and the number of topics. For the first iteration, LDA randomly assigns each word to a topic. With each iteration over the abstracts, the algorithm updates the probability that each word belongs to a particular topic. We found that the topics did not substantially change beyond 200 iterations. For the number of topics, we initially reviewed sets of 20, 35, 50, and 75. We found that the set of 20 failed to capture some of the topics in the set of 35; however, several of the topics in the set of 35 appeared to be very similar to one another, and the topics began to break down as the number further increased. Therefore, we decided to pursue a 30-topic model for labeling. To interpret the topics, two of the authors independently reviewed the 10 words with the highest probability of co-occurring in each topic, and the 20 abstracts represented by the highest proportions of each topic. Overall, the authors had a strong level of agreement, and resolved any disagreements about topic labels through discussion.

We include both MeSH term and topic modeling analyses because they provide different and complementary ways of understanding how actionability is conceptualized in biomedical literature. Because MeSH terms are annotated by humans, the terms associated with each abstract will likely be more complete and conform to a well-designed hierarchy of terms that categorize millions of scientific papers. Thus, our MeSH term analysis allows us to clearly examine which fields and subfields have taken up the concept of actionability in different years. In comparison, the topic modeling results we provide are more complete because they include abstracts that have not yet been annotated with MeSH terms (21% of our sample) and are more specific to the actual words and phrases most associated with different types of abstracts mentioning actionability. This more granular analysis allows us to see in more detail how different technologies, concepts, and practices are related to the framework of actionability. Utilizing both methods also serves as a form of replication to ensure that both analyses underline the same main findings.

## Results

As shown in Figure 1, the first abstract to mention “actionable” or “actionability” in the PubMed database occurred in 1978. Qualitative manual review of the first 100 abstracts that include actionability as a keyword (published between 1978 and 2006) suggests that the term was first used in the literature to reference a legal definition of actionable (i.e., when there are legal grounds to file a lawsuit). Then, the concept slowly started proliferating in meaning, often referenced as a metric used to process information, aid in policy or priority-setting, or provide quality assurance measures. Mentions of actionability did not appear frequently until approximately 2006—the first year in our sample to reach 20 abstracts. By 2012, the number of abstracts mentioning actionability grew to over 100, and has continued rising significantly since that point, reaching over 1800 abstracts in 2022. Thus, we find two main inflection points where actionability gained traction within biomedical science: the mid 2000s, and the early 2010s. In the Conclusion, we provide hypotheses for why actionability may have taken root in these periods.

**Figure 1.**
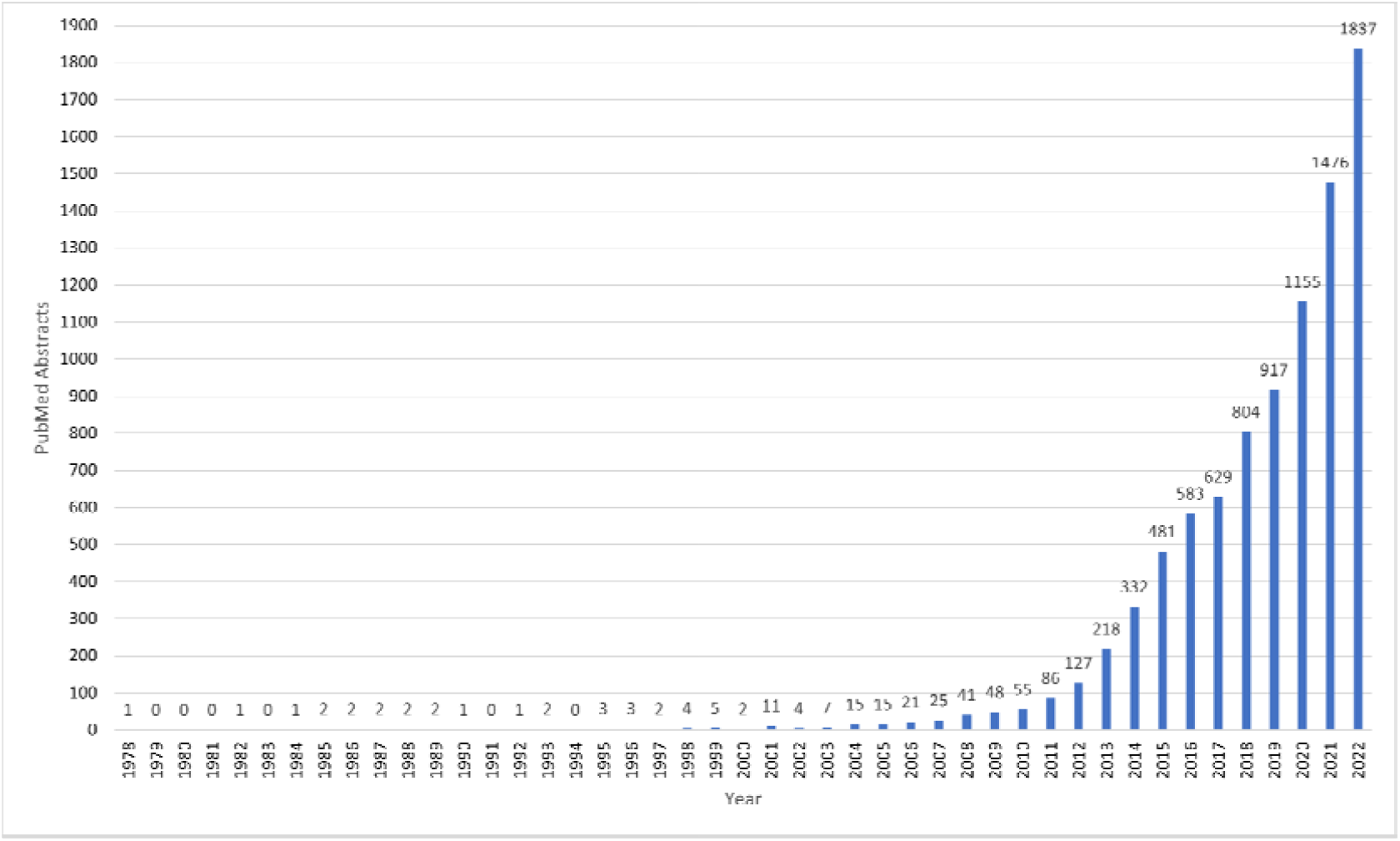
Total number of abstracts mentioning “actionable” or “actionability” indexed in PubMed, per year. This figure excludes incomplete data from 2023.

### MeSH terms

To understand how actionability was taken up by different fields and subfields, we analyzed the MeSH terms used to categorize articles archived in PubMed. As of December 2022, MeSH terms were available for 7074 (79%) of the 8943 abstracts in PubMed that mentioned “actionable” or “actionability.” There were 6583 unique MeSH terms associated with these 7074 abstracts. Figure 2 presents the proportion of abstracts associated with common, select MeSH terms, per year. This figure visualizes a few key trends. First, two of the three most common MeSH terms, both peaking in mid 2010s, are related to the field of genetics/genomics: “Mutation” and “High-Throughput Nucleotide Sequencing.” Other common MeSH terms from this field include “Genomics,” “Precision Medicine,” and “Genetic Predisposition to Disease. The other field represented most prominently in the MeSH terms is oncology, including: “Neoplasms,” “Biomarkers, Tumor,” and “Antineoplastic Combined Chemotherapy Protocols.” Some common terms combine these two fields, such as “Molecular Targeted Therapy.” Figure 3 provides a more streamlined visualization of the MeSH term analysis, confirming that the quick uptake of actionability in published literature in the early 2010s was driven primarily by its increased use in the fields of genetics/genomics and oncology (neoplasms/cancer).

**Figure 2.**
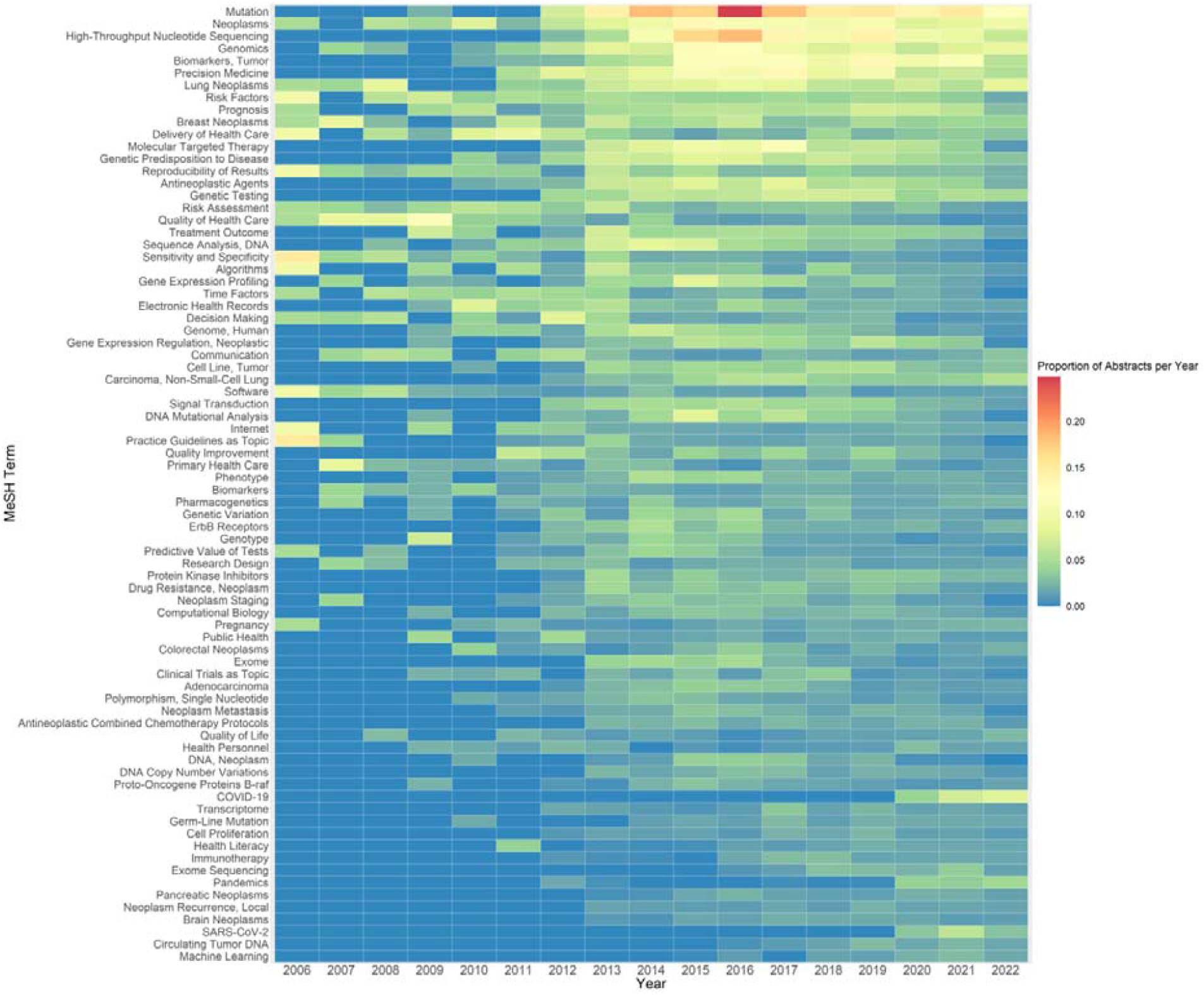
Proportion of abstracts associated with select MeSH terms, per year. This figure includes 78 (76%) of the 102 MeSH terms that occurred at least 100 times among the 7074 abstracts in our dataset, excluding 24 terms determined not to be informative for this study, such as those indicating demographics (e.g., *female*, middle-aged, *United States*) or study designs (e.g., *retrospective studies, surveys, and questionnaires, qualitative research*). This figure excludes the years prior to 2006, when there were fewer than 20 abstracts published per year that mentioned “actionable” or “actionability.” Sixteen of the terms listed were first introduced as MeSH terms between 2006-2022 and thus were not represented in the abstracts until their introduction (such as MeSH term “COVID-19,” introduced in 2020). For full details on when these 16 terms were introduced, see Supplementary Table 1.

**Figure 3.**
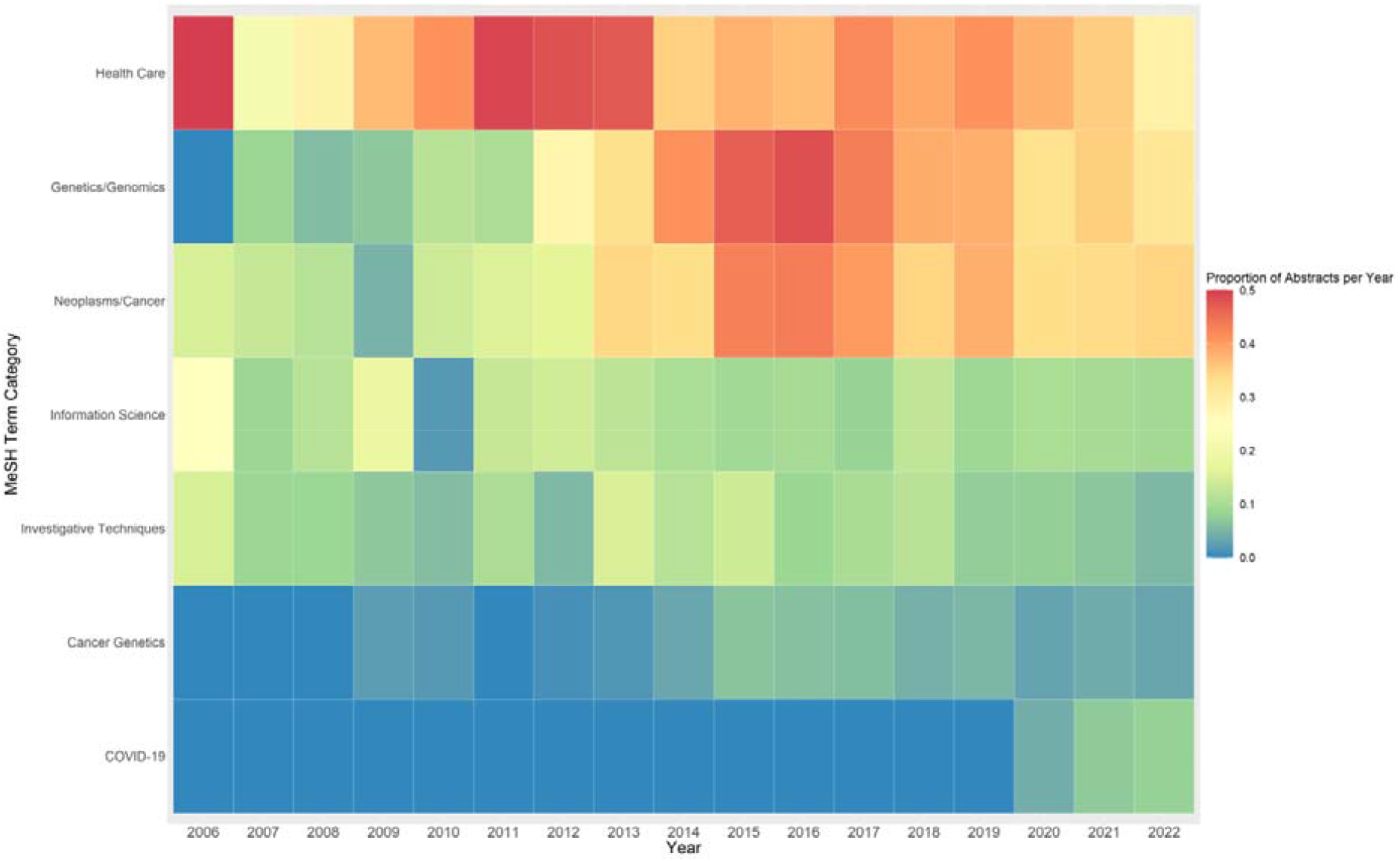
Proportion of abstracts associated with select high-level MeSH terms, per year. Figure 3 groups the MeSH terms in Figure 2 into 7 higher-level categories, based in part on the MeSH hierarchy, excluding 3 of the MeSH terms in Figure 1 that we determined did not belong neatly to one of these categories: *Signal Transduction, Biomarkers*, and *Pregnancy*.

### Topic modeling

In addition to our analysis of MeSH terms, we used topic modeling to confirm and provide additional detail on the emergence and proliferation of actionability in published literature. Table 1 presents the 10 words with the highest probability of co-occurring in each of the 30 topics, and the label that was manually assigned to each topic based on these words. Four of the topics were similar enough that we reviewed an additional 10 words to help differentiate their labels: “Cancer Treatment,” “Cancer Genetics,” “Clinical Genetic Sequencing,” and “Return of Genetic Test Results.” Table 1 also presents both the total number of abstracts represented by each topic and the number of abstracts that are most represented by each topic. For example, while the “Community Health” topic is represented in the largest total number of abstracts, the “Cancer Treatment” topic is the most salient topic in the largest number of abstracts. Figure 4 presents the proportion of abstracts represented by each topic per year, between 2006-2022.

**Table 1.**
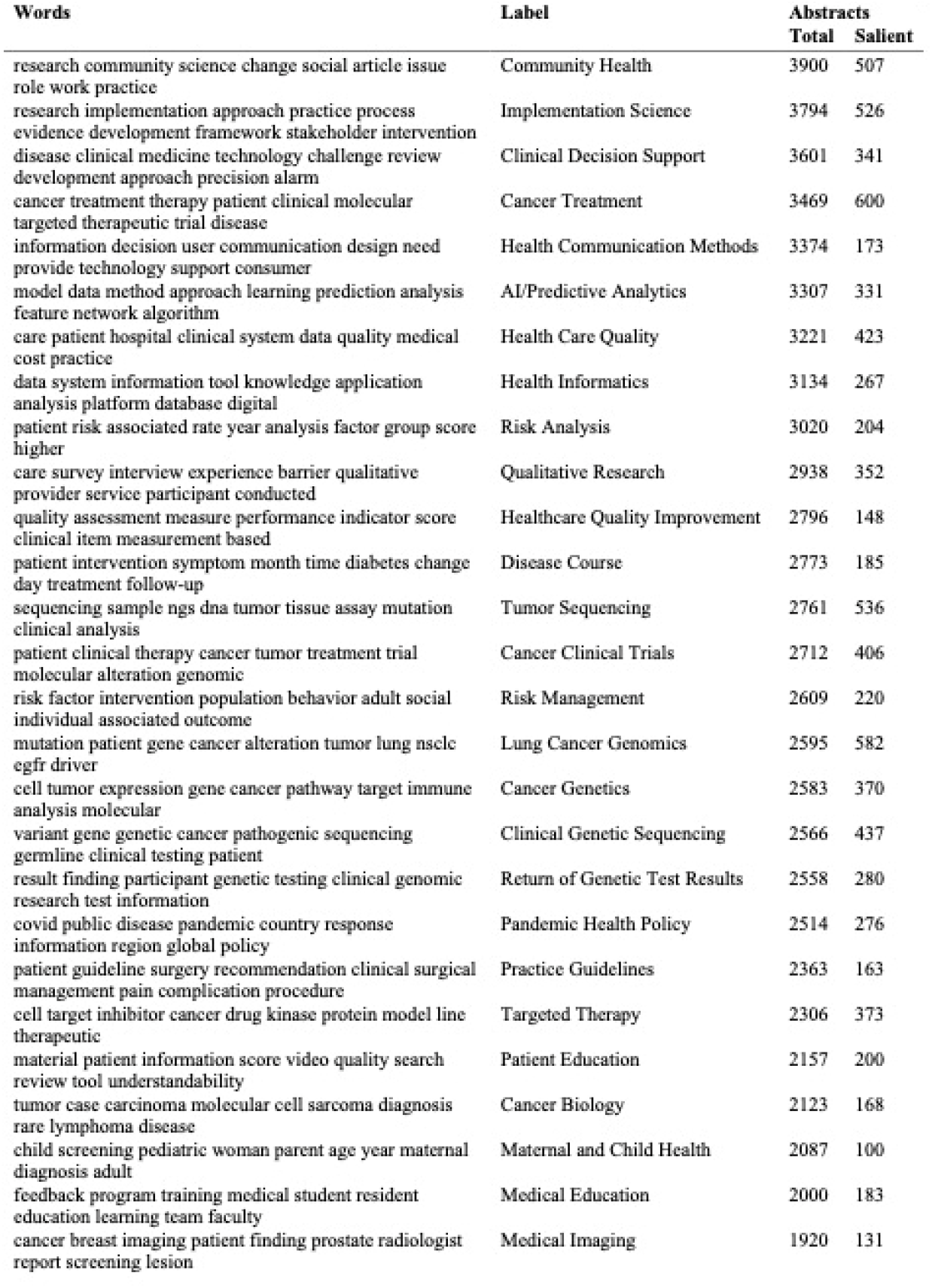
Topic modeling labels, words with highest probability of co-occurrence per topic, and abstracts represented per topic.

**Figure 4.**
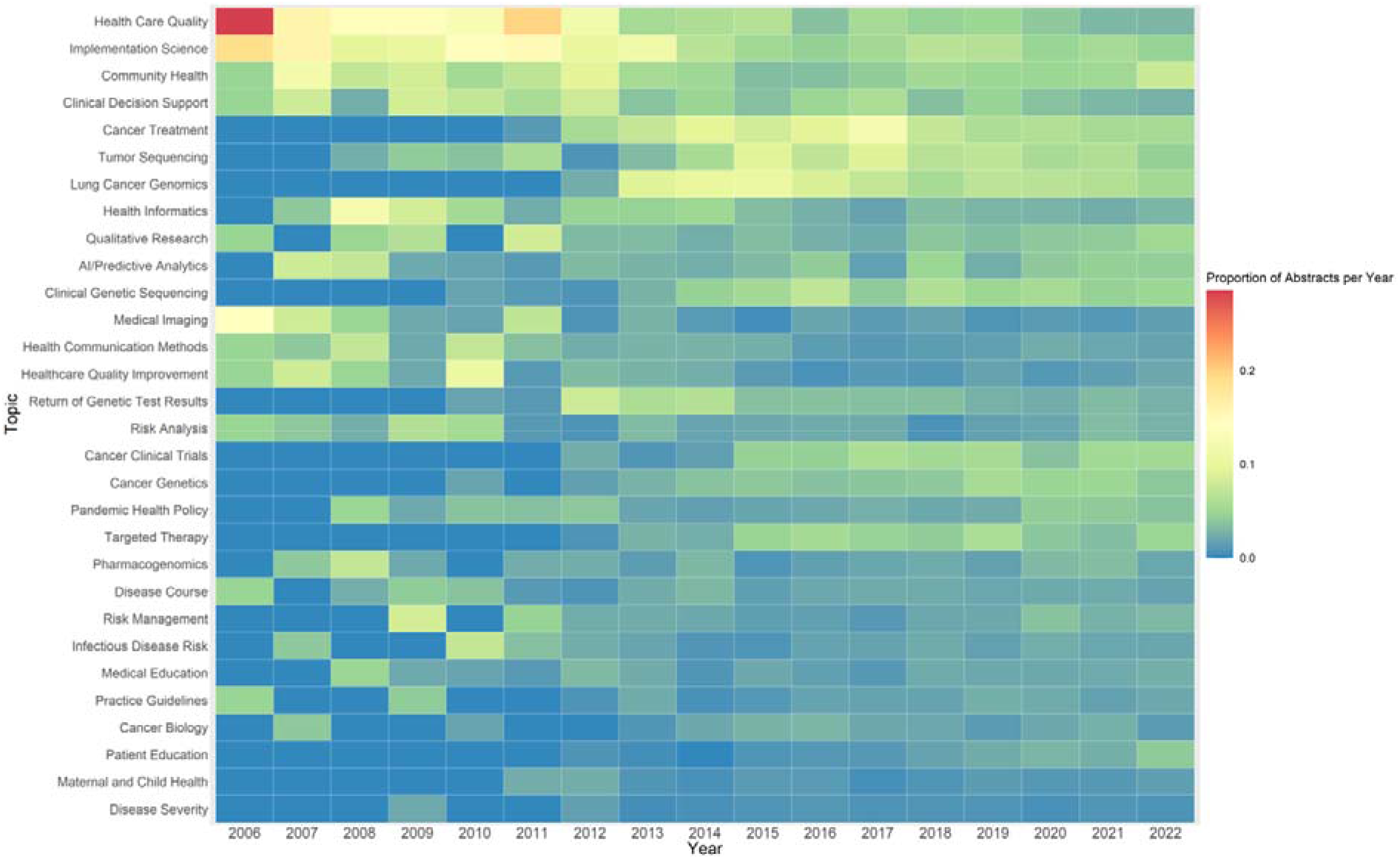
Proportion of abstracts represented by each topic, per year. Each abstract is counted only once towards the topic most represented in that abstract, and the sum of the proportions of abstracts per year is equal to 1.

Figure 4 demonstrates that much of the early discussions of actionability related to health care quality, implementation, clinical decision support, or health communication/education, and that these topics remain well-represented through the entire dataset. At the same time, Figure 4 shows that the precipitous rise in use of actionability as a framework was again driven by genetics and oncology, with topics such as “Tumor Sequencing,” “Lung Cancer Genomics,” “Clinical Genetic Sequencing,” and “Targeted Therapy” appearing more frequently starting in the mid-2010s. Thus, two main themes emerge from this data in the mid-2010s: the use of genetic technologies such as tumor sequencing in cancer treatment, and identifying genetic predisposition to diseases (often cancers).

## Conclusions

In our examination of the concept of actionability in published literature, we find that actionability seems to be discussed frequently in two contexts: (1) tumor sequencing and targeted therapies for cancer treatment, and (2) identifying genetic predispositions to disease. At the same time, actionability was also commonly discussed in healthcare implementation, quality improvement, and communication/education. Based on these findings, we theorize that there are two main areas in biomedical research where the framework of actionability provides value for researchers and clinicians: (1) enacting new policy or procedure in an effective way, and (2) acting on data generated from new forms of testing, such as next-generation sequencing. While references to actionability in the context of policy and procedure remained more stable across our dataset, interest in actionability as a means to understand new technology has spiked in the past 10 years.

The concept of actionability first gained traction in the mid-2000s and proliferated in the 2010s. The two fields primarily driving this proliferation are genetics/genomics and oncology. Based on this data, we suggest a few hypotheses to understand these two inflection points. Most saliently, the completion of the Human Genome Project in 2003 sparked tremendous innovation in translational medicine as researchers sought to incorporate genetic data and insights into clinical care. The framework of actionability may have helped to make sense of this new resource and technology. The early 2000s also brought advances in cancer treatment and research, including the first tyrosine kinase inhibitor (a targeted therapy) approved by the Food and Drug Administration (FDA) in 2001 (Zhong et al. 2021) and the Cancer Genome Atlas Program, launched by the National Cancer Institute and the National Human Genome Research Institute in 2006 (Hutter and Zenklusen 2018). With these new developments, a critical question became: how can researchers identify actionable molecular targets for new cancer therapies (Carr et al. 2016; Mateo et al. 2018)? We hypothesize that these innovations in genetic sequencing and new knowledge of the molecular basis of cancer likely prompted increased discussions of actionability in the mid-2000s.

Then, the early 2010s marked the most precipitous rise in discussions of actionability, prompting our analysis of what occurred in this period that could generate this interest. We hypothesize that decreases in the cost of genetic sequencing could be the driving force behind this uptake of actionability in published literature. For example, Illumina had produced the first $1000 genome by 2014, marking tremendous progress in reducing the cost of sequencing. We also note that the number of articles represented in our dataset seems inversely proportional to the cost of genome sequencing (National Human Genome Research Institute 2021), indicating that technological advances such as next generation sequencing may have fostered discussions of the actionability of genetic information. Because genomic sequencing became more cost-effective for a range of medical applications, professional societies like the American College of Medical Genetics and Genomics (ACMG) introduced guidelines for the return of incidental genetic findings, widely embracing the concept of “actionable genes” by 2016 (Kalia et al. 2017).

Our findings also lead us to assess what made the framework of actionability particularly valuable for genetics and oncology. First, we suggest that actionability is a framework that is well-suited for genetics and other data-intensive fields seeking to make sense of tremendous amounts of data. In comparison to fields that tend to collect data with a specific hypothesis in mind, genetics often works in reverse, trying to find key insights and hypotheses from existing biobanks of health data. In this scenario, the value of a research finding could be assessed via its actionability—does this information seem likely to lead to action or change in clinical treatment or care? In oncology, the framework of actionability could be appealing because researchers are often trying to identify which kinds of tumor mutations are actually relevant and may lead to an effective treatment option. As other researchers have demonstrated, there are both tremendous research efforts underway to identify new actionable molecular targets for cancer therapies and ongoing discussion of what should count as an actionable mutation (Carr et al. 2016).

Despite these rationales for the appeal of actionability as a framework to assess the value of health technologies, we also urge caution to consider the drawbacks of conceptualizing value in this way. Actionability drives our attention primarily toward whether a test or piece of information can lead to action, not whether that action has proven benefits. In comparison to other ways of evaluating the value of a new intervention, like clinical validity or utility, actionability may aim toward the overuse of interventions with limited or untested effectiveness. As the Choosing Wisely initiative of the ABIM Foundation has shown, overuse of interventions is still a common problem in medicine, leading to unnecessary costs and potential negative health outcomes from unneeded treatments (Cassel and Guest 2012). Thus, while actionability can be a valuable way to identify new innovations in translational medicine, evaluations of clinical efficacy will require other frameworks of knowledge production.

By tracing the social history of actionability in biomedicine through computational analyses of published literature, we provide new insights on the proliferation of actionability as a framework to assess value that adds to a large body of literature on the production of knowledge and evidence in translational medicine. Our finding that the popularity of actionability as a framework is primarily driven by the fields of genetics and oncology suggests that additional research is needed to understand the implications of that framework more fully for these two fields. More generally, bioethics analyses of the underlying values and assumptions embedded in actionability as a framework for knowledge production will be crucial as clinicians and researchers continue to face difficult questions about how to sort through tremendous amounts of data to generate knowledge that can have a real impact on patient health.

## Data Availability

All data produced in the present study are available upon reasonable request to the authors.

## Acknowledgments

This work was supported by the National Human Genome Research Institute (R00HG010905). The content is solely the responsibility of the authors and does not necessarily represent the official views of the National Institutes of Health. The authors thank Ivan Flores for contributing to data collection, and José Agustín Gutiérrez Gómez for contributing to data analysis.

---

**Supplementary Table 1.** Year of introduction for MeSH terms that first appear between 2006-2022.

| **MeSH Term** | **Year of Introduction** |
| --- | --- |
| Circulating Tumor DNA | 2016 |
| COVID-19 | 2020 |
| DNA Copy Number Variations | 2010 |
| Electronic Health Records | 2009 |
| ErbB Receptors | 2012 |
| Exome | 2012 |
| Exome Sequencing | 2016 |
| Health Literacy | 2010 |
| High-Throughput Nucleotide Sequencing | 2011 |
| Machine Learning | 2015 |
| Molecular Targeted Therapy | 2011 |
| Pandemics | 2011 |
| Precision Medicine | 2010 |
| Quality Improvement | 2011 |
| SARS-CoV-2 | 2020 |
| Transcriptome | 2012 |

